# The health impact of long COVID during the 2021-2022 Omicron wave in Australia: a quantitative burden of disease study

**DOI:** 10.1101/2022.08.01.22278219

**Authors:** Samantha Howe, Joshua Szanyi, Tony Blakely

**Affiliations:** Population Interventions Unit, Centre for Epidemiology and Biostatistics, Melbourne School of Population and Global Health, The University of Melbourne, Carlton, VIC, 3053

## Abstract

**Background:** Long COVID symptoms occur for a proportion of acute COVID-19 survivors, with reduced risk among the vaccinated, and for Omicron compared to Delta variant infections. The health loss attributed to pre-Omicron long COVID has previously been estimated using only a few major symptoms.

**Methods:** The years lived with disability (YLDs) due to long COVID in Australia during the 2021-2022 Omicron BA.1/BA.2 wave were calculated using inputs from previously published case-control, cross-sectional, or cohort studies examining the prevalence and duration of individual long COVID symptoms. This estimated health loss was compared with acute SARS-CoV-2 infection YLDs and years of life lost (YLLs) from SARS-CoV-2. The sum of these three components equals COVID-19 disability-adjusted life years (DALYs); this was compared to DALYs from other diseases.

**Results:** 5200 (95% uncertainty interval [UI] 2200-8300) YLDs were attributable to long COVID and 1800 (95% UI 1100-2600) to acute SARS-CoV-2 infection, suggesting long COVID caused 74% of the overall YLDs from SARS-CoV-2 infections in the BA.1/BA.2 wave. Total DALYs attributable to SARS-CoV-2 were 50 900 (95% UI 21 000-80 900), 2.4% of expected DALYs for all diseases in the same period.

**Conclusion:** This study provides a comprehensive approach to estimating the morbidity due to long COVID. Improved data on long COVID symptoms will improve the accuracy of these estimates. As data accumulates on SARS-CoV-2 infection sequelae (e.g., increased cardiovascular disease rates), total health loss is likely to be higher than estimated in this study. Nevertheless, this study demonstrates that long COVID requires consideration in pandemic policy planning given it is responsible for the majority of direct SARS-CoV-2 morbidity, including during an Omicron wave in a highly vaccinated population.

**Key messages:**

- Our study is the first to comprehensively estimate long COVID morbidity using its individual symptoms, during Australia’s 2021-2022 Omicron wave.
- We show that long COVID contributed to almost three-quarters of the non-fatal health loss resulting from Omicron infections in this period.
- Long COVID contributes to a substantial proportion of direct COVID-19 morbidity, even in a highly vaccinated population during an Omicron wave. It should therefore be more explicitly considered in future pandemic policymaking.
- Our method of estimating long COVID morbidity has explicable differences to existing long COVID burden of disease approaches and may provide a more accurate estimate of the morbidity attributable to long COVID.

## Introduction

A post-acute phase of SARS-CoV-2 infection, commonly termed long COVID, occurs among some individuals following acute infection. Long COVID describes the persistence and/or emergence of a heterogeneous group of symptoms at least 12 weeks after acute infection.^1^ There is no current consensus on the symptom profile that specifically characterises long COVID, with a wide range of symptoms being reported across multiple organ systems, including cardiopulmonary, neurological, and musculoskeletal systems.^2^ While some studies report symptom clustering within individuals, the frequency and significance of this remains unclear and individuals most commonly report experiencing one or two symptoms.^3,4^ The aetiology of long COVID is proposed to be related to continued immune activation and persistence of the virus in the various organ systems infected during the acute period.^5^ Numerous risk factors have been identified, including a propensity towards an autoimmune response, female sex, co-morbidities such as Type 2 Diabetes Mellitus, and a more severe acute infection.^4,6^ Importantly, COVID-19 vaccination has been found to reduce the risk of long COVID.^7,8^

Given ongoing high rates of SARS-CoV-2 transmission globally, it is important to quantify the full health impact of SARS-CoV-2 infection, including its longer-term consequences. Recent burden of disease studies have quantified long COVID by treating it as a single outcome, utilising the Global Burden of Disease (GBD) study health state of ‘post-acute consequences’ from other respiratory illnesses, or chronic fatigue syndrome, to approximate long COVID severity.^9-11^ While these health states have some overlap with documented long COVID symptoms, they do not acknowledge the full breadth of symptoms linked to long COVID, or the heterogeneity of symptoms reported between individuals.^2,9^

A lack of high-quality evidence regarding the prevalence and duration of symptoms hampers quantification of long COVID burden. Many studies of long COVID lack an appropriate control group or are subject to other methodological issues. Of particular concern is selection bias from self-selection into studies by those experiencing ongoing symptoms and loss-to-follow up by those no longer symptomatic. These biases likely inflate estimates of long COVID occurrence. Conversely, studies with insufficient follow-up time may not capture the full burden of long COVID symptoms, which are often relapsing and remitting in nature.^2^ In addition to these design issues, the majority of published long COVID research has been conducted in cohorts of unvaccinated, pre-Omicron variant infected cases. To accurately quantify the impact of long COVID in highly vaccinated populations during Omicron waves, differences in the risk of long COVID by vaccination and variant need incorporating.

This paper addressed the following research questions: firstly, what is the extent of the morbidity attributable to long COVID resulting from Omicron variant SARS-CoV-2 infections, quantified ‘bottom-up’ from occurrence rates of each symptom and its severity and duration? Secondly, what proportion of total years lived with disability (YLDs) and disability-adjusted life years (DALYs) accumulated during the 2021-2022 Omicron wave in Australia were due to long COVID, and how does this health loss compare to other major causes of health loss in Australia?

## Methods

### Morbidity calculations

In this study, the impact of long COVID in the Australian population is measured with a burden of disease approach. Disability-Adjusted Life Years are a standard metric of disease burden and encompass both morbidity (as YLDs), and mortality (as years of life lost [YLL]). YLDs are the focus of this paper, as long COVID is treated as a collection of non-fatal symptoms; however overall DALYs combining both acute and long COVID are also measured.

Long COVID morbidity was calculated as that expected per symptomatic SARS-CoV-2 infected person:

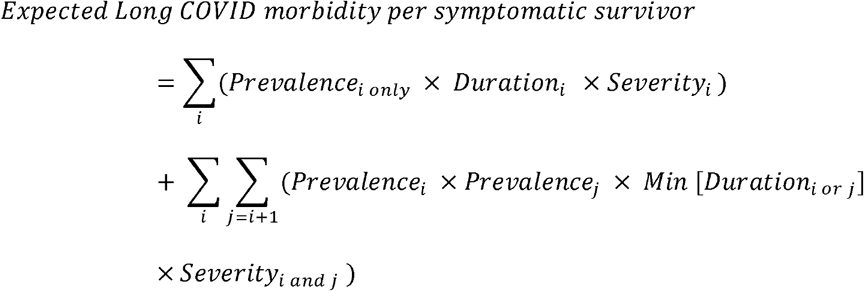

where *i* and *j* index each possible symptom, and *Prevalence*_*i only*_ is the prevalence of symptom *i* minus the sum of the joint occurrence of symptom *i* with each other symptom (assuming symptom occurrence is independent). The prevalence of each symptom was calculated as a risk difference in symptom frequency between SARS-CoV-2 survivors and SARS-CoV-2-negative controls, extracted from previous research (see Supplementary Table 1 and Supplementary Table 2, available as Supplementary data at *IJE* Online).^3,12-14^ Only symptoms found to occur more frequently in COVID-19 cases compared to COVID-negative controls, in the controlled literature, were included (presented in Table 1). The severity of each symptom was quantified as a disability weight (DW) taken from the GBD study ^15^ for the matching health state, and best matches otherwise (e.g. DWs of other sensory conditions have been used as proxies for dysosmia and dysgeusia). *Severity*_*i only*_ was calculated as ^16^:

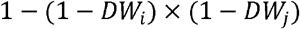

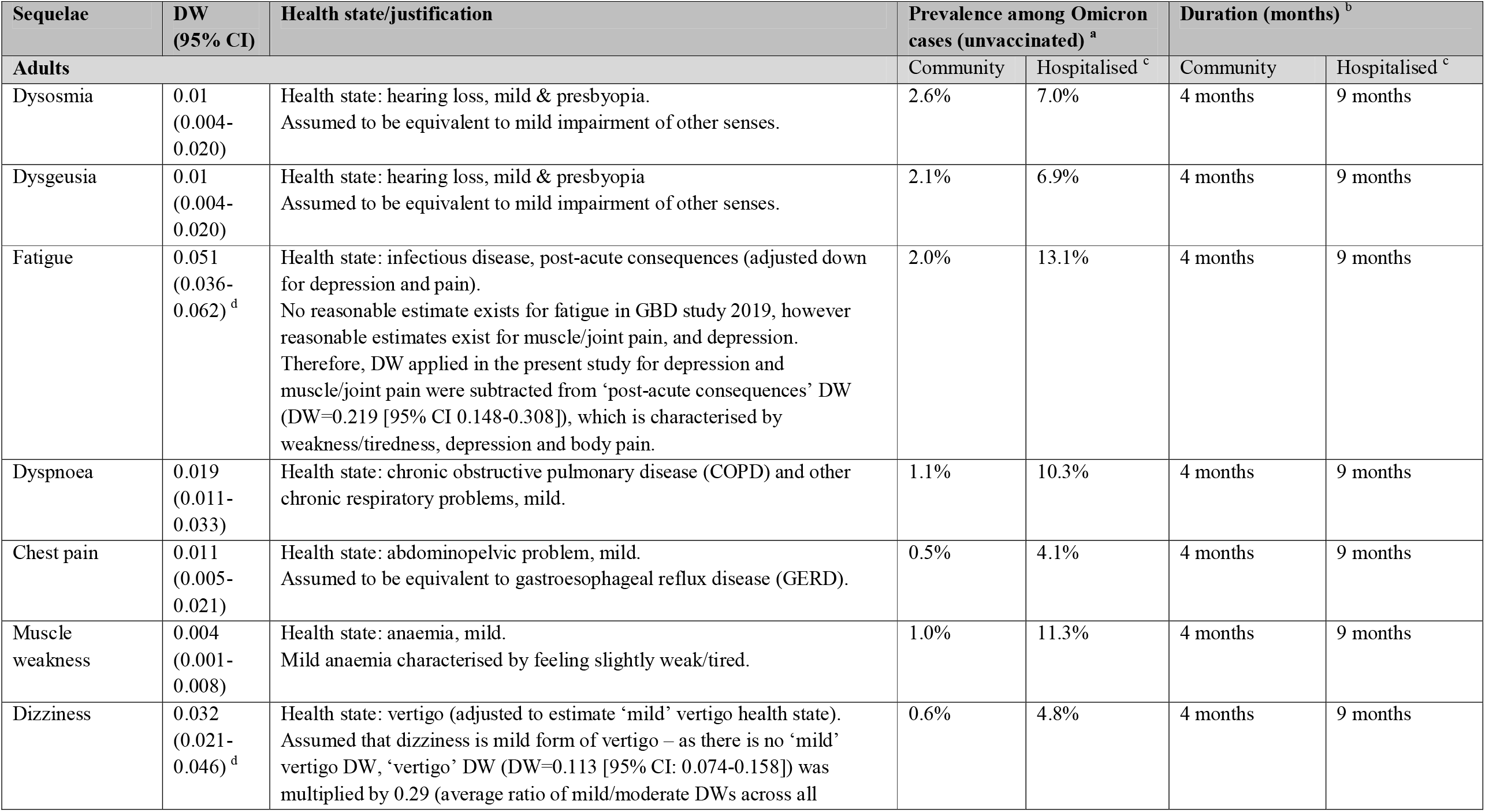
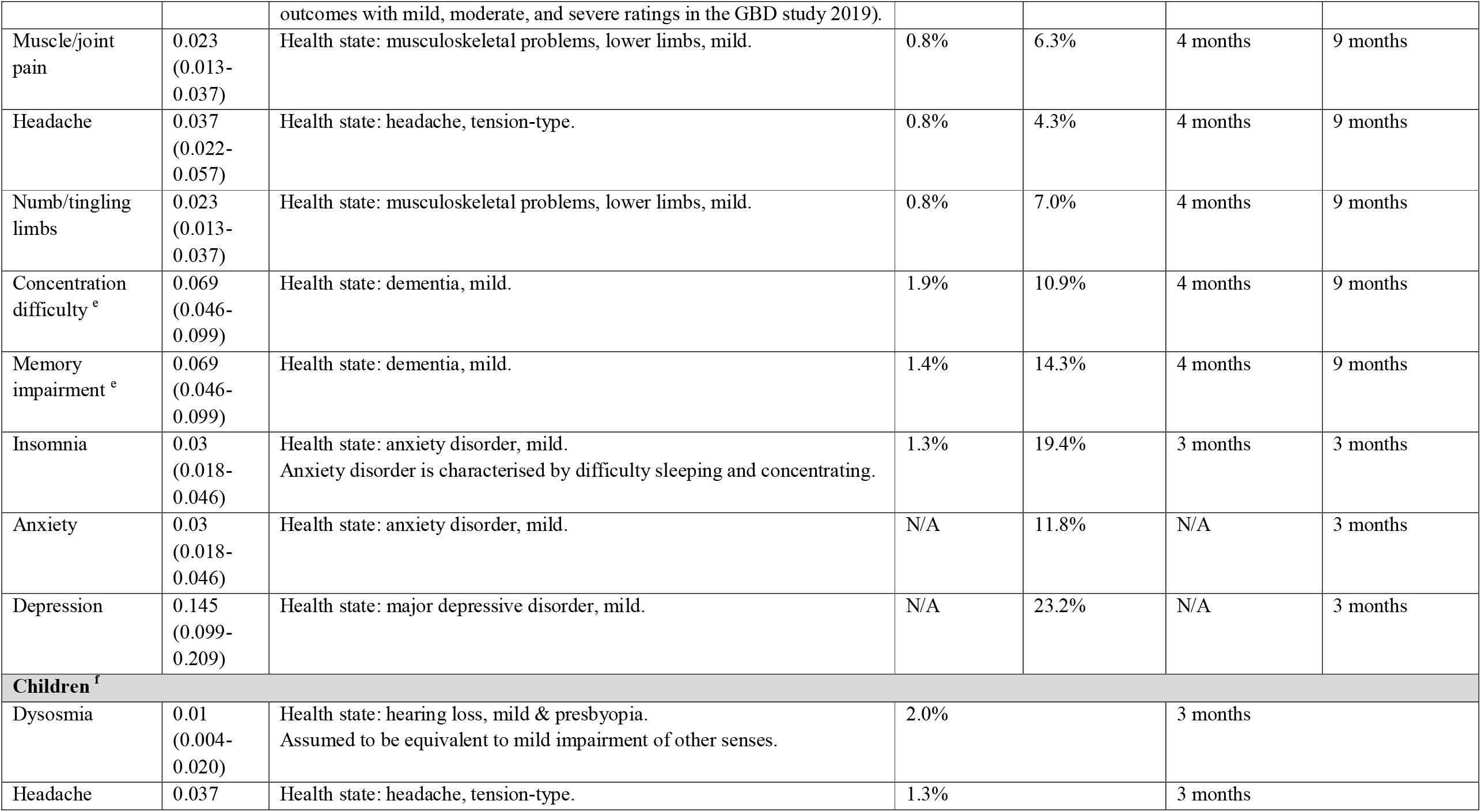
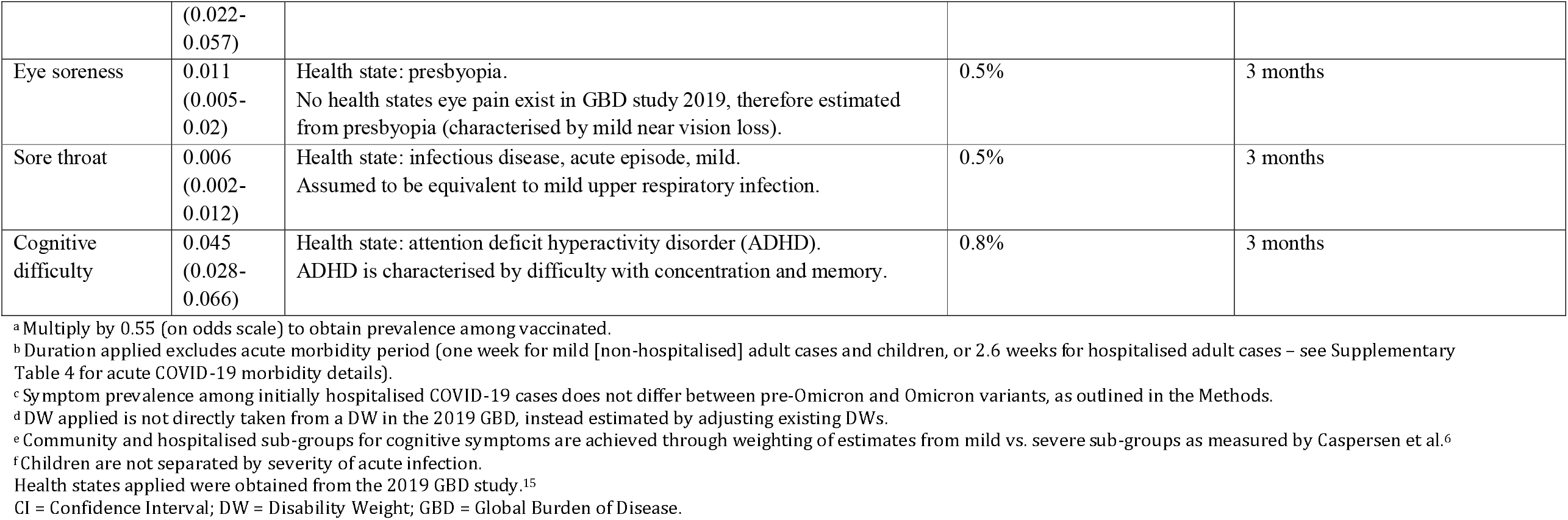

It was assumed that only initially symptomatic patients (during the acute illness period) are at risk of long COVID – the risk of long COVID among asymptomatic patients has been found to be low or nil compared to symptomatic patients. ^17-19^ We also assumed that calculations including all three-way (or higher) combinations of symptoms would make negligible difference to morbidity estimates – given that most symptom prevalence estimates are less than 10% (Table 1), and joint prevalence calculations are multiplicative, the joint prevalence of three or more symptoms using this method would be very low. Additionally, it is uncommon to experience more than two symptoms).^12^

The above ‘base case’ prevalence calculations were for unvaccinated individuals infected with a pre-Omicron variant of SARS-CoV-2, and were split into three sub-groups: adult community cases, adult hospitalised cases, and children (0-17 years, any acute disease severity).^3,12-14^ Long COVID symptom prevalence among previously hospitalised adults is approximately twice that among community cases.^12^ Symptom prevalence is also lower among children compared to adults.^14^ Duration of each symptom (from one week post-infection for mild/moderate cases, and approximately 2.5 weeks post-infection for those hospitalised) was applied based on recent findings by Wulf Hansen et al., who reported a median duration of symptoms for community infections of four months, and 8.9 months for previously hospitalised cases.^20^ Exceptions to this have been applied to psychological symptoms, for which a shorter duration was used, and similarly for children (see Table 1).^13,21^

These base case prevalences were then multiplied by an odds ratio (OR) of 0.55 to approximate symptom prevalence among vaccinated cases based on findings from two studies that found reduced odds of symptoms following the acute infection by 49% and 41%, for those who had at least two COVID-19 vaccines compared to one/no vaccines.^7,8^ Vaccination post-infection has been found to have a limited effect on long COVID occurrence – therefore, only vaccination prior to infection was considered.^22,23^ Next, prevalence estimates were further multiplied by an OR of 0.25 based on an estimate of the reduction in prevalence of any symptoms at least four weeks post-infection for Omicron variant compared to Delta variant infections in a vaccinated cohort in the United Kingdom.^24^ It was assumed that this association remains beyond 12 weeks post-infection. The exception to the latter multiplier was adults hospitalised with an Omicron infection (the previously mentioned study was conducted using a cohort of primarily community cases); we assumed that once a case is severe enough to be hospitalised, there is no difference in resulting long COVID compared to pre-Omicron variants. A flow diagram depicting the above method to cross walk prevalence data from pre-Omicron, unvaccinated cases to Omicron, vaccinated cases is presented in Figure 1.

**Figure 1:**
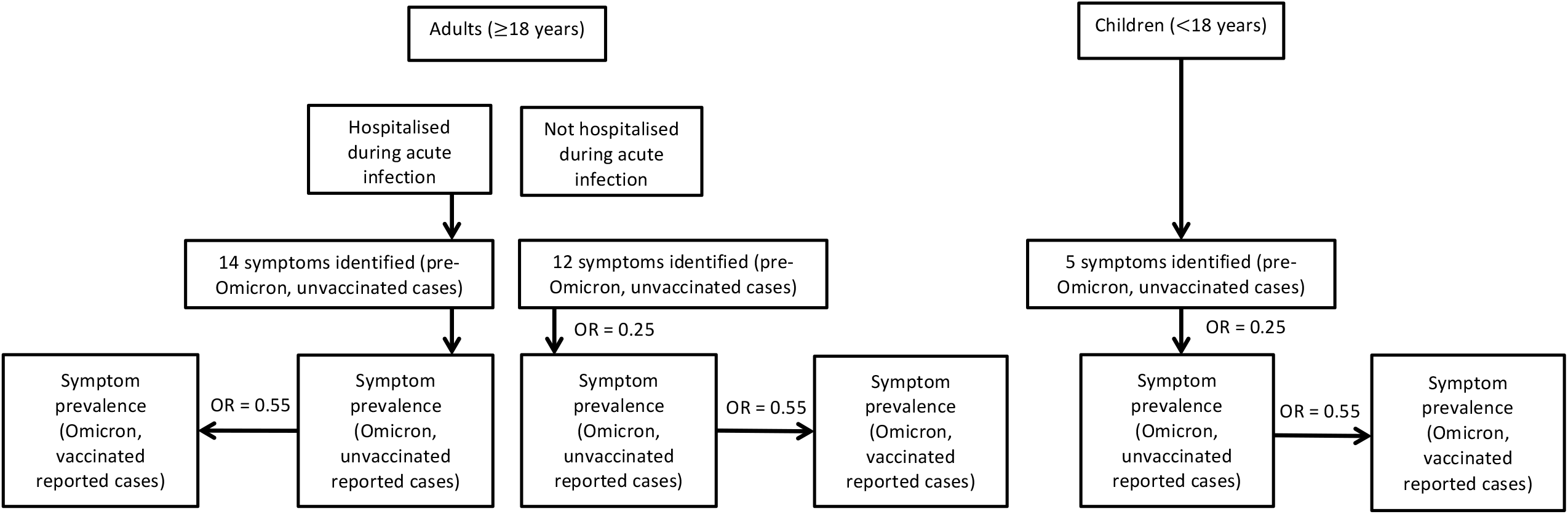
Flow diagram – prevalence crosswalk method. OR=0.25 indicates reduced odds of long COVID among Omicron-infected cases compared to Delta-infections - applied to not hospitalised (i.e., community) adult cases and children. OR=0.55 indicates reduced odds of long COVD among vaccinated (at least 2 COVID-19 vaccines) compared to unvaccinated (<2 vaccines) cases – applied to all vaccinated sub-groups. OR = Odds ratio

Uncertainty was included using a standard deviation (SD) of +/- 20%, applied to base case long COVID morbidity estimates. This method approximated the 95% uncertainty in base case estimates achieved using a more comprehensive approach, where variance in duration, severity and prevalence were each calculated to estimate total variance. For morbidity calculations for vaccinated and Omicron-infected populations, we used a SD of +/- 30% of the expected value to reflect additional uncertainty.

The DW, estimated prevalence, and average duration of each identified long COVID symptom among Omicron infected, unvaccinated cases, along with the uncertainty applied, are presented in Table 1. The prevalence among vaccinated cases is that shown in Table 1 multiplied by 0.55. The estimated prevalence and duration of each symptom among base cases (i.e., unvaccinated, pre-Omicron infections) is presented in Supplementary Table 3, available as Supplementary data at *IJE* online.

### Application to the Omicron wave and health burden comparison

Long COVID YLDs were calculated as the expected long COVID morbidity per symptomatic case multiplied by the total number of symptomatic infections (treated as equivalent to notified cases) during the four months of the Omicron BA.1/BA.2 wave in Australia, defined here as December 10^th^ 2021 to April 9^th^ 2022.

The YLDs from acute SARS-CoV-2 infection were estimated based on a previously published method by Blakely et al. ^25^, updated here to be specific to Omicron-variant infections. Acute COVID-19 morbidity is sub-divided into community cases and hospitalised cases that are either ward only or include an ICU admission – approximately 8% of cases in hospital have been estimated as requiring ICU admission during the first four months of the Omicron wave.^26^ Symptom duration estimates for hospitalised patients were based on findings from a New South Wales (NSW) study by Tobin et al.^27^, which estimated hospital stay duration during December (at the start of the Omicron wave). These duration estimates were weighted across the three age categories presented by Tobin et al.^27^ (0-39 year olds, 40-69 year olds and 70+ year olds) based on the proportion hospitalised in each age group. A percentage split of 72% mild and 28% moderate acute illness for non-hospitalised cases was based on findings by Menni et al. ^28^, which indicated that the odds of moderate severity illness was reduced by approximately 44% for Omicron compared to Delta infections. Previous estimates used by Blakely et al.^25^ for pre-Omicron variant infections utilised a 50/50 split for the proportions of moderate vs. mild community cases. Acute COVID-19 morbidity inputs are presented in Supplementary Table 4, available as Supplementary data at *IJE* online.

YLLs due to SARS-CoV-2 deaths were estimated using standard burden of disease methods ^29^, multiplying reported deaths due to COVID-19 with the average remaining life expectancy of those who died (with reference life expectancy by sex and age obtained from the 2019 GBD, retrieved from the Institute for Health Metrics and Evaluation [IHME] results tool).^30,31^

YLL and YLD calculations were by strata of age (<18-year-olds vs. ≥18-year-olds), hospitalisation status (as described above) and vaccination – national level data was not available at the time of writing for vaccination status of COVID-19 cases, therefore data from NSW was applied.^32^

The total DALYs from SARS-CoV-2 infection in these four months were calculated as the sum of the above three components. For the purposes of this paper, we assigned the long COVID YLDs to the SARS-CoV-2 infections occurring in the four-month window.

For comparison, we took the YLDs and DALYs in Australia for other diseases from GBD 2019 (adjusted to reflect the 2021 population [mid-year])^31,33^, dividing by three to make them equivalent to the four-month duration of the BA.1/BA.2 wave.

### Sensitivity analyses

One-way sensitivity analyses were conducted on base case morbidity estimates, separately varying the prevalence, severity and duration components by their 95% uncertainty intervals (UI). Additionally, each base case estimate was varied for the adult cohorts by the symptoms extracted from different source studies. This included varying the prevalence, duration and severity components together by their respective 95% UI, separately for: physical symptoms obtained from Sørensen et al.^12^; psychological symptoms from Caspersen et al.^3^; and cognitive symptoms from Magnúsdóttir et al.^13^).

An extreme scenario analysis was also conducted in which the OR for Omicron compared to pre-Omicron infections (OR=0.25) was applied to the hospitalised patient group, as applied for community cases only in the main analysis.

## Results

Some 4.87 million COVID-19 cases were notified in Australia during the first four months of the Omicron wave, with approximately 35 500 hospitalisations and 3463 deaths. ^26,30,34,35^ An estimated 61% of notified cases were vaccinated at the time of infection (noting that approximately 90% of 16+ year olds in the population were fully vaccinated by 10 December 2021).^32,36^ Reported cases and deaths are shown in Supplementary Table 5 and Supplementary Table 6, available as Supplementary data at *IJE* online.

The overall morbidity (i.e. YLDs) resulting from infections in this period, including acute and long COVID is shown in Figure 2 (with YLDs by sub-group presented numerically in Table 2). Long COVID accounted for approximately 74% of the overall non-fatal COVID-19 health loss among notified cases during the first 4 months of the Australian Omicron wave, at 5200 YLDs (95% uncertainty interval [UI] 2200-8300). The majority of long COVID YLDs come from community cases, with the highest number in the vaccinated community adult cases sub-strata (2100 YLDs, 95% UI 900-3500) given that this group represents the greatest proportion of notified cases (Supplementary Table 5, available as Supplementary data at *IJE* online). Per person COVID morbidity estimates, which show the non-fatal health loss due to long COVID per notified COVID-19 case are shown for each sub-group in Supplementary Table 7, available as Supplementary data at *IJE* online.

**Table 2:** COVID-19 YLDs resulting directly from Omicron cases during the first 4-months of the Omicron wave, December 10^th^ 2021-April 9^th^ 2022.

| Population group | YLD (95% UI <sup>a</sup>) |  |  |
| --- | --- | --- | --- |
|  | Acute COVID-19 | Long COVID | Total |
| **Hospitalised adults (unvaccinated) <sup>b</sup>** | 43 (26-60) | 431 (178-684) | 498 (205-791) |
| **Hospitalised adults (vaccinated) <sup>b</sup>** | 89 (54-124) | 375 (154-595) | 488 (201-774) |
| **Non-hospitalised adults (unvaccinated)** | 425 (258-591) | 1967 (810-3123) | 2391 (985-3798) |
| **Non-hospitalised adults (vaccinated)** | 867 (527-1207) | 2197 (905-3488) | 3064 (1262-4865) |
| **Children (unvaccinated)** | 255 (155-355) | 171 (70-271) | 426 (176-677) |
| **Children (vaccinated)** | 157 (95-218) | 59 (24-94) | 216 (89-343) |
| **Total** | **1835 (1116-2555)** | **5247 (2162-8333)** | **7035 (2898-11 171)** |

<sup>a</sup> 95% UI estimated using +/- 30% standard deviation for long COVID and total YLDs, +/- 20% standard deviation for acute COVID-19 YLDs.
<sup>b</sup> For acute COVID-19 morbidity, hospitalised group included ICU and ward only calculations.
Note: figures presented in the text have been rounded from values in this table.
UI= Uncertainty Interval; YLD= Years Lived with Disability

**Figure 2:**
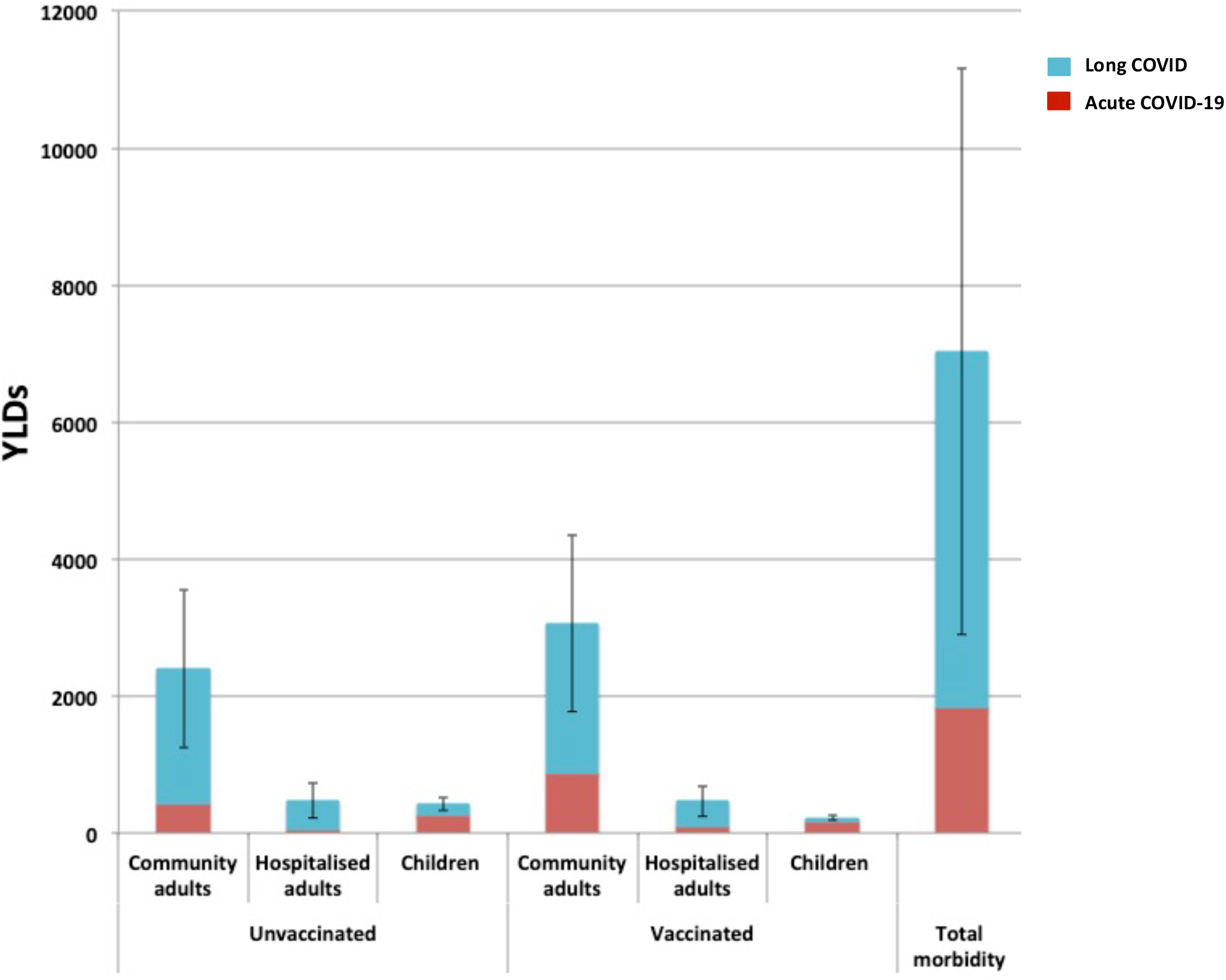
COVID-19 YLDs resulting directly from Omicron cases during the first 4-months of the Omicron wave, December 10^th^ 2021-April 9^th^ 2022. 95% uncertainty intervals are shown for long COVID YLD estimates, measured with +/- 30% standard deviation. Total morbidity (rightmost bar) is estimated at 7000 YLDs. YLDs= Years Lived with Disability

The overall DALYs resulting from reported COVID-19 infections in the four-month period was estimated at 50 900 (95% UI 21 000-80 900), of which long COVID contributed 10.3% (compared to the acute COVID-19 morbidity contribution of 3.6%). The remaining DALYs resulted from acute COVID-19 mortality.

The leading 25 causes of morbidity in Australia ranked by YLDs are presented in Figure 3A along with the estimated YLDs resulting from reported COVID-19 cases during the first four months of the Omicron wave. Total COVID-19 morbidity is ranked 24^th^, comparable to the non-fatal health loss resulting from chronic kidney disease.

**Figure 3:**
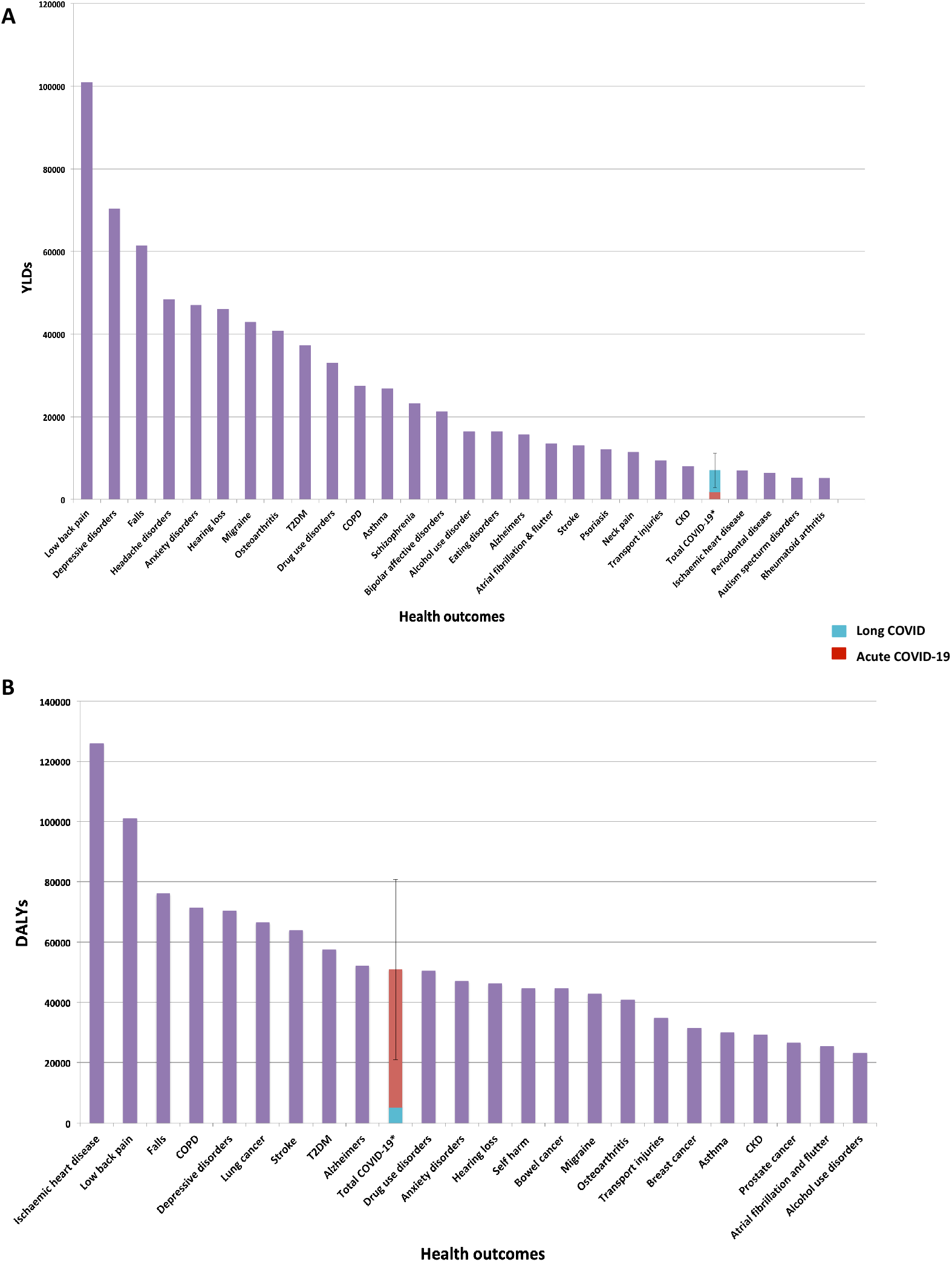
Burden of disease comparison during the first 4 months of the Omicron wave (December 10^th^ 2021-April 9^th^ 2022), Australia. **Panel A:** Comparison of the YLDs due to COVID-19 separated as long COVID (blue) and acute COVID-19 (red), to other outcomes (purple). Total COVID-19 YLDs = 7000 (95% uncertainty interval 2900-11 200). **Panel B:** Comparison of the DALYs due to COVID-19 separated as long COVID (blue) and acute COVID-19 (red), to other outcomes (purple). Total COVID-19 DALYs = 50 900 (95% uncertainty interval 21 000-80 900). Note that COVID-19 YLDs and DALYs include the future morbidity resulting from long COVID for these cases. 95% uncertainty intervals are shown for COVID-19 DALY and YLD estimates (+/- 30% standard deviation). YLDs and DALYs for other outcomes are estimated from the 2019 GBD study, updated to the population size as of June 2021, and subsequently divided by three to estimate health loss over a four-month period.^15^ COPD= Chronic Obstructive Pulmonary Disease; CKD= Chronic Kidney Disease; T2DM= Type 2 Diabetes Mellitus; YLD= Years Lived with Disability. *Total COVID-19 YLDS/DALYS include acute COVID-19 and long COVID.

The leading 25 causes of health loss in Australia ranked by DALYs, are presented in Figure 3B along with the estimated DALYs resulting from the first four months of the COVID-19 Omicron wave. The DALYs resulting from COVID-19 cases are estimated to be 2.4% of all expected DALY loss in the four months, and rank 10^th^ among conditions (between Alzheimer’s and other dementias, and drug use disorders). The 95% UI indicates that the rank could in fact be as high as 3^rd^ (between low back pain and falls) or as low as 24^th^ (below alcohol use disorders).

## Discussion

We estimated that long COVID contributed to approximately 74% of the non-fatal health loss (i.e., YLDs) resulting from reported COVID-19 infections in the first four months of the Omicron BA.1/BA.2 wave in Australia. Given that morbidity was calculated using notified cases, and some unreported cases will have been symptomatic, this may be an underestimate. The total YLDs for acute and long COVID combined were estimated as comparable to YLDs caused by chronic kidney disease and ischaemic heart disease (Figure 3A). The overall COVID-19 disease burden, YLDs plus years of life lost, was estimated at 50 900 DALYs (95% UI 21 000-80 900) over the initial four months of the Omicron wave, the 10^th^ highest cause of DALYs in this four-month period in Australia and approximately 2.4% of all health loss (Figure 3B).

Our per person morbidity estimates for long COVID show that for a vaccinated adult case who did not require hospitalisation during their acute infection (i.e. the majority of cases in the first four months of the Omicron wave in Australia), 0.09% of a healthy year of life is lost due to long COVID (equivalent to one third of a day of healthy life lost). It is important to note that the morbidity loss for someone actually with long COVID would be much greater than this, as this estimate (and those shown in Supplementary Table 7, available as Supplementary data at *IJE* online) gives the health loss for any surviving notified COVID-19 case.

A strength of our study is allowing for the lower incidence of long COVID among vaccinated people, and with Omicron versus pre-Omicron variants; to our knowledge, previous studies have not made both allowances. Our approach also quantifies long COVID morbidity across multiple patient sub-groups and includes a complete symptom profile rather than treating long COVID as a single outcome.

A recent study by Wulf Hanson et al. and Institute for Health Metrics and Evaluation (IHME) collaborators estimated the proportion of SARS-CoV-2 infections developing long COVID, separated into three symptom groups, for SARS-CoV-2 infections in 2020 and 2021.^20^ Their method involved measuring the prevalence of each symptom individually as well as the prevalence of each symptom pair and all three symptoms together. Using this method, then applying odds ratios for long COVID for Omicron compared to pre-Omicron, and vaccination status, our morbidity estimate observing the equivalent symptom groups is within 20% of the resulting estimate from the IHME method (as shown in Supplementary Table 8, available as Supplementary data at *IJE* online).^20^ This indicates that our assumption of independence is valid for burden estimation of long COVID.

Our approach does not provide an overall estimate of long COVID occurrence rate, given that we define long COVID as a heterogeneous group of symptoms, rather than a single outcome. The utility in our approach is in the use of severity and duration estimates specific to each symptom, to more accurately estimate the morbidity attributed to long COVID in a population. The use of single health states, such as ‘post-acute consequences’ (as currently recommended by the European Burden of Disease Network)^37^ assumes that the average long COVID sufferer has all these symptoms included within the applied health state. It also ignores a large proportion of symptoms that have been identified among long COVID sufferers. In Supplementary Table 9 (available as Supplementary data at *IJE* online), we compare our morbidity estimate with those from other COVID-19 burden of disease studies that have incorporated long COVID. Long COVID parameters published by the Australian Institute of Health and Welfare (AIHW) ^10^, and adjusted by us to reflect vaccinated, Omicron infections, result in an estimate approximately two-thirds of our finding, likely due to the reduced duration applied (91 days) and the use of a single health state (post-acute consequences; DW=0.219). Three other non-Australian publications, which applied an even shorter duration of 28 days, with a single long COVID health state, produced estimates approximately one-third of our morbidity estimate.^38-40^ These differences highlight the strength of our paper, which accounts for a wider range of long COVID symptoms, and a longer average duration of symptoms reflecting that documented in the existing literature. Our approach also accounts for the difference in risk of long COVID symptoms by acute COVID-19 severity, and age, which these publications have not considered. We postulate that our ‘bottom up’ approach provides a more accurate measure of morbidity resulting from long COVID.

Our study has limitations. Long COVID is an area of rapidly emerging and evolving research, currently characterised by limited high-quality literature. As such, there is a high level of uncertainty in our findings (e.g., our ‘cross-walk’ of pre-Omicron unvaccinated data to Omicron vaccinated cases). Additionally, while the studies utilised for our prevalence estimates were included based on an assessment of bias and methodological quality, prevalence may still be overestimated, due to the likely direction of bias to over-estimation in long COVID studies. A recently published prospective cohort study that controlled for symptoms present prior to COVID-19 diagnosis, found a slightly reduced occurrence of symptoms among COVID-positive cases compared to controls than our base case analysis.^41^ While these estimates were not able to be used in this paper, given that the study did not stratify data by age and severity of acute infection, as more data becomes available on the prevalence of symptoms among strata Omicron-infected cases our estimates can be updated. Further stratification of patient groups may also help to improve the accuracy of our current framework, for example by increasing the number of age strata and including sex. There is an increased risk of long COVID among older age groups and females ^42^, however, data on symptom prevalence was not available allow for this in our analysis in addition to the variables we included.

Given the lack of data on the occurrence of long COVID symptoms among previously hospitalised Omicron cases, and the known association between acute COVID-19 severity and long COVID risk,^12^ we assumed no difference in long COVID morbidity between the hospitalised base case group (i.e. pre-Omicron infections) and Omicron hospitalised cases. In a sensitivity analysis, we tested an ‘extreme’ for this sub-group, applying the odds ratio (OR=0.25) used for cross walking of community pre-Omicron to community Omicron cases, to the hospitalised group.^24^ While this reduced the morbidity for the hospitalised patient group by an average of 81.5% compared to the main analysis, overall long COVID morbidity was only reduced by 12% compared to the main analysis, to 4600 YLDs (95% UI 1900-7200) (see Supplementary Table 10, available as Supplementary data at *IJE* online). This estimate largely overlaps with that from our primary analysis, however, it will still be important to conduct further research into long COVID among hospitalised Omicron cases in the future, including consideration of those requiring ICU treatment vs. ward treatment only (which was not possible in this study given a lack of data available).

Additional one-way sensitivity analyses, presented as Tornado plots, showed that the majority of the overall uncertainty in long COVID morbidity in our study was due to uncertainty in the morbidity severity estimates for symptoms (i.e. disability weights), as opposed to uncertainty in symptom frequency and duration (Supplementary Figure 1, available as Supplementary data at *IJE* online). This is of concern, given that the majority of DWs applied were made by estimation from other health states ^15^, and points to the need for research to better quantify the severity of each long COVID symptom.

It is important to note that many people dying of COVID-19 have co-morbidities, so their true YLLs will be less than that measured using standard DALY methods, which assumes those dying of COVID-19 have the mortality rate specified in the reference life table. It is also important to note that our study focuses on health loss from acute and long COVID; SARS-CoV-2 infection has also been associated with persistent organ damage, and an increased risk of chronic conditions including Type 2 Diabetes Mellitus and cardiovascular disease, particularly among those who had a severe acute infection.^43-47^ Longer-term research will be required in the future to determine the extent that SARS-CoV-2 infection causes these sequelae, which will lead to the total burden of SARS-CoV-2 being (perhaps considerably) greater than that quantified in this study.

## Conclusion

Our approach for estimating the morbidity attributable to long COVID likely provides a more accurate measure of long COVID burden compared to that of existing burden of disease studies. Whilst the prevalence of long COVID symptoms is less now among highly vaccinated populations with Omicron than it was for pre-Omicron variants among unvaccinated populations, long COVID still contributes substantial health loss when summed over all infections. Our findings therefore highlight the need to factor in long COVID-related health loss when making policy decisions.

## Data Availability

All data/sources used for quantitative analysis can be made available by authors on reasonable request.

## Ethical approval

This study did not require ethical approval.

## Author contributions

Conceptualisation: S.H., J.S., T.B. Literature review: S.H. Formal analysis: S.H., T.B. Writing – original draft: S.H., J.S., T.B. Writing – review & editing: S.H., J.S., T.B.

## Data availability

The data underlying this article will be shared on reasonable request to the corresponding author

## Supplementary data

Supplementary data are available at *IJE* online.

## Acknowledgements

Expert knowledge was kindly provided by Professor John D. Potter (Massey University, Wellington, Fred Hutchinson Cancer Research Centre, Seattle, Washington and University of Washington, Seattle, Washington).

