## Supplementary Material for "The health impact of long COVID during the 2021-2022 Omicron wave in Australia: a quantitative burden of disease study"

**Supplementary data**

### **Supplementary Table 1: Literature search strategy**

| **Database searches** | PubMed, Scopus, Medline (Ovid), Web of Science  **Search date:** 17/03/22 |
| --- | --- |
| **Search terms** | "long COVID" OR "post-acute COVID" OR "post acute COVID" OR "post-acute sequelae" OR "post acute sequelae" OR “post-COVID syndrome” OR “post COVID syndrome” |
| **Non-database/other searches** | **Pre-print database:** MedRvix  **COVID-19 repositories:** The Lancet COVID-19 resource centre, the BMJ Coronavirus hub  Reference lists from relevant studies identified in above searches  **Search date:** ongoing |
| **Inclusion criteria** | **Study type:** cohort studies, case-control studies, cross-sectional studies  **Date published:** 2020-present  **Timeframe:** measurements taken at least 90 days post-COVID diagnosis (to meet long COVID definition)  **Study population:** COVID-positive cases confirmed by  PCR/antigen/serology testing compared to COVID-negative controls confirmed by PCR/antigen/serology testing    **Outcome:** any new onset sequelae occurring post-COVID diagnosis |
| **Exclusion criteria** | **Study type:** review/commentary, randomised controlled trials  **Date published:** prior to 2020  **Timeframe:** measurements only taken less than 90 days post-COVID diagnosis  **Study population:** no control group or inappropriate control group that is not representative of general population (e.g. hospitalised due to other respiratory viruses), non-population based study sample (e.g. health care workers), loss to follow-up greater than 50%  **Outcome:** sequelae present prior to COVID-19 infection, outcomes measured using electronic health records only (only captures outcomes/symptoms reported to a doctor/health service, so does not capture the full breadth of symptoms occurring post-acute infection) |

### **Supplementary Table 2: Studies utilised for long COVID morbidity estimates**

| **Study** | **Study features** | **Symptoms measured** | **Potential biases/methodological concerns** |
| --- | --- | --- | --- |
| **Sørensen et al. (2022, preprint) ^12^**  Denmark  Cross-sectional study | **Sample source/recruitment:**  Population based – individuals who had a PCR test from Sep 2020- Apr 2021 invited via nationally utilised electronic communication system  **Sub-group:**  Adults (18+), community & hospitalised acute infection  **Number of participants (% Female):**  61,002 cases (58.7% Female)  91,878 controls (62.8% Female)  **Follow up/time frame:**  6-12 months (data collected for cases 6, 9 and 12-months post-infection) | Dysosmia  Dysgeusia  Fatigue  Dyspnoea  Muscle weakness  Sleeping/numb limbs  Muscle/joint pain  Headache  Dizziness  Chest pain  Hot flushes/sweat ^a^  Reduced appetite ^a^  Red runny eyes ^a^  Abdominal pain ^a^  Chills ^a^  Nausea ^a^  Diarrhoea ^a^  Fever ^a^  Cough  Runny nose  Sore throat | **Selection bias:**   - Low risk of selection bias: similar response rate between cases and controls (35.6% & 35.5% invited individuals participated)   **Information bias:**   - Misclassification of the exposure unlikely: PCR tests confirmed for all cases and controls using electronic health record system - Misclassification of the outcome possible: self-reported symptoms with single question survey responses for each symptom, with low/moderate risk of recall bias as participants only asked to state what symptoms they have experienced in the last 14 days (health problems reported to occur prior to COVID-19 diagnosis additionally excluded) - Exception to survey questions is for psychological & cognitive symptoms: participants were asked to state any diagnoses received in the last 6-12 months related to these symptoms (also not stratified by hospital status during acute infection) – therefore, these symptoms not included   **Methodological issues:**   - Comparisons cannot be directly made between time points, as they represent different cases |
| **Magnúsdóttir et al. (2022)** ^13^  Multi-country cohort study & meta-analysis (Denmark, Estonia, Iceland, Norway, Sweden, United Kingdom) | **Sample source/recruitment:**  Different strategies used across cohorts  **Sub-group:**  Adults (18+), community & hospitalised acute infection (in supplementary material only)  **Number of participants (% female):**  9979 cases (67.9% female)  237,270 controls (61.7% female)  **Follow up/time frame:**  2-16 months (differed across cohorts) | Depression ^b^  Anxiety ^b^  COVID-related PTSD  Poor sleep (insomnia) | **Selection bias:**   - Recruitment methods varied across cohorts: some cohorts were created prior to the pandemic, while others specifically recruited those testing for COVID-19 - Greater proportion females compared to males   **Information bias:**   - Misclassification of the exposure possible: all cohorts relied on self-report of positive PCR/antibody test for COVID-19 - Misclassification of the outcome is unlikely: validated health questionnaires utilised, in a prospective cohort format (therefore, low risk of recall bias)   **Methodological issues:**   - Small hospitalised sub-group (main analysis was between those bedridden during acute illness vs. not bedridden – hospitalisation comparison presented only in supplementary material as not all cohorts measured hospitalisation status during acute infection) |
| **Caspersen et al. (2021)** ^3^  Norway  Cross-sectional survey within ongoing cohort study | **Sample source/recruitment:**  Existing population-based cohort study participants (pre-pandemic)  **Sub-group:**  Adults (18+), mild and severe acute infection  **Number of participants (% female):**  774 cases (170 in wave 1 cohort, 583 in wave 2 cohort) (58% female)  72,953 controls (59.3% female)  **Follow up/time frame:**  1-12 months (1-6 months post-infection cohort from wave 2, 11-12 months post-COVID cohort from wave 1) | Fatigue  Hair loss  Palpitations  Joint pain  Muscle pain  Altered smell/taste  Skin rash  Chest pain  Cough  Headache  Dyspnea  Heart palpitations  Reduced lung function  Concentration difficulty  Memory impairment  Dizziness  Anxiety  Depression ^c^  Mood swings ^c^  Sleep difficulty  Myocarditis  Hair loss  Kidney disease  Fever | **Selection bias:**   - Low/moderate risk of selection bias: used pre-existing population based cohort (i.e. recruitment not specific to COVID-19); case response may have been over-represented by those experiencing symptoms (however, no major baseline characteristic differences between cases and controls)   **Information bias:**   - Misclassification of the exposure unlikely: confirmed PCR test for all cases and controls via online national registry - Misclassification of the outcome possible: self-reported symptoms with single question survey responses for each symptom, with potential for recall bias (however, participants only asked to state what symptoms they are currently experiencing, and any symptoms that were reported as being experienced for a period of >12 months/longer time frame than time since COVID-19 diagnosis were excluded)   **Methodological issues:**   - Small hospitalised sub-group - Comparisons cannot be directly made between time points, as they represent different cases |
| **Behnood et al. (2022)** ^14^  Systematic review of 5 controlled studies | **Number of studies:**  3-5 studies included for meta-analysis of each symptom  **Sub-group:**  Children (0-18 years), any acute infection severity  **Follow up/time frame:**  4 weeks-6 months across studies included in meta-analyses | Cognitive difficulties  Headache  Loss of sense of smell  Sore throat  Sore eyes  Abdominal pain  Cough  Fatigue  Muscle pain  Insomnia  Diarrhoea  Fever  Dizziness  Dyspnoea | **Selection bias:**   - Each study had sample from different populations, with differing recruitment methods - Heterogeneity ranged from 0.02-98% across meta-analyses   **Information bias:**   - Misclassification of the exposure possible: one of the studies included used self-report of COVID-19 - Misclassification of the outcome possible: all studies were cohort studies so lower risk of recall bias, but some studies included used only proxy-report by parents/guardians of participants   **Methodological issues:**   - One of the studies in included had a timeframe of only 4 weeks, the other only 8 weeks post-infection (i.e. cannot be defined as long COVID) (Zavala et al., Molteni et al.)^5,6^ |

^a^ Symptoms with risk difference between all cases and controls of less than 2% are not included in analysis (Sørensen et al.).^12^

^b^ Symptoms may only be present in hospitalised/severe cases, not community/mild cases (Magnúsdóttir et al.).^13^

^c^ Symptoms may only have greater frequency in cases compared to controlled for the 1-6 month post-COVID group (wave 2) not the 11-12 month post-COVID group (wave 1) (Caspersen et al.).^3^

Symptoms occurring at a greater frequency in cases compared to controls are underlined (considering only those symptoms where the confidence interval does not cross the null value of 0).

### **Supplementary Table 3: Long COVID symptom profile, with prevalence and duration, estimated among pre-Omicron unvaccinated cases**

| **Symptom** | **Prevalence among pre-Omicron cases (unvaccinated) ^a^** | **Duration ^b^** |
| --- | --- | --- |
| **Adults community cases** | | |
| **Dysosmia** | 10.4% | 4 months |
| **Dysgeusia** | 8.2% | 4 months |
| **Fatigue** | 7.8% | 4 months |
| **Dyspnoea** | 4.2% | 4 months |
| **Chest pain** | 1.8% | 4 months |
| **Muscle weakness** | 4.1% | 4 months |
| **Dizziness** | 2.3% | 4 months |
| **Muscle/joint pain** | 3.2% | 4 months |
| **Headache** | 3.0% | 4 months |
| **Numb limbs** | 3.2% | 4 months |
| **Concentration difficulty ^c^** | 7.6% | 4 months |
| **Memory impairment ^c^** | 5.5% | 4 months |
| **Insomnia** | 5.3% | 3 months |
| **Adult hospitalised cases** | | |
| **Dysosmia** | 7.0% | 9 months |
| **Dysgeusia** | 6.9% | 9 months |
| **Fatigue** | 13.1% | 9 months |
| **Dyspnoea** | 10.3% | 9 months |
| **Chest pain** | 4.1% | 9 months |
| **Muscle weakness** | 11.3% | 9 months |
| **Dizziness** | 4.8% | 9 months |
| **Muscle/joint pain** | 6.3% | 9 months |
| **Headache** | 4.3% | 9 months |
| **Numb limbs** | 7.0% | 9 months |
| **Concentration difficulty ^c^** | 10.9% | 9 months |
| **Memory impairment ^c^** | 14.3% | 9 months |
| **Insomnia** | 19.4% | 3 months |
| **Anxiety** | 11.8% | 3 months |
| **Depression** | 23.2% | 3 months |
| **Children ^d^** | |  |
| **Dysosmia** | 8.0% | 3 months |
| **Headache** | 5.0% | 3 months |
| **Eye soreness** | 2.0% | 3 months |
| **Sore throat** | 2.0% | 3 months |
| **Cognitive difficulties** | 3.0% | 3 months |

^a^ Prevalence estimates presented here are for the population of unvaccinated, pre-Omicron COVID-19 cases. Prevalence estimates for adult community cases and children (but not adults hospitalised) are multiplied by 0.25 (odds scale) to reflected expected long COVID prevalence resulting from Omicron variant infections, as shown in the main analysis (Table 2). To achieve the prevalence among vaccinated cases, all estimates are multiplied by 0.55 (odds scale).

^b^ Duration applied excludes acute morbidity period (one week for mild [non-hospitalised] adult cases and children, or 2.6 weeks for hospitalised adult cases – see Supplementary Table 4 for acute COVID-19 morbidity details).

^c^ Community and hospitalised sub-groups for cognitive symptoms are achieved through weighting of estimates from mild vs. severe sub-groups as measured by Caspersen et al. ^3^

^d^ Children are not separated by severity of acute infection.

### **Supplementary Table 4: acute COVID-19 morbidity parameters**

| **Category** | **Duration** **in days** | **GBD health state** | **DW (95% CI)** | **Morbidity** |
| --- | --- | --- | --- | --- |
| Admitted hospital, including ICU | 7 | Moderate acute infection | 0.051 (0.032-0.074) | 0.00098 |
|  | 3.41 | Severe acute infection (for hospital stay) | 0·133 (0.088–0.190) | 0.00124 |
|  | 4.3 | COPD (for ICU) ^a^ | 0.408 (0.273-0.556) | 0.00481 |
|  | 14 | Return to baseline health | Linearly to 0 from COPD DW | 0.00782 |
|  | **Total** | | | 0.0149 |
| Admitted to hospital, ward only | 7 | Moderate acute infection | 0.051 (0.032-0.074) | 0.00098 |
|  | 3.41 | Severe acute infection (for hospital stay) | 0.133 (0.088–0.190) | 0.00124 |
|  | 7 | Return to baseline health | Linearly to 0 from severe acute infection DW | 0.00128 |
|  | **Total** | | | 0.0035 |
| Symptomatic, non-hospitalised | 6.87 | Moderate acute infection (=28%) | 0.051 (0.032-0.074) | 0.00096 |
|  | 6.87 | Mild acute infection (=72%) | 0.006 (0.002–0.012) | 0.00011 |
|  | **Total** | | | 0.00035 |

^a^ Severe COPD used as the health state to estimate ARDS (as seen for severe COVID-19 requiring ICU admission), for which no DW exists in the GBD study. ^15^

Adapted from Blakely et al. and Tobin et al.^25,27^

ARDS= Acute Respiratory Distress Syndrome; CI= confidence interval; COPD= Chronic Obstructive Pulmonary Disease; DW= Disability Weight; GBD= Global Burden of Disease; ICU= Intensive Care Unit.

### **Supplementary Table 5: Omicron wave cases**

| **Case groups** | **Non-hospitalised** | | **Hospitalised ^a^** | |
| --- | --- | --- | --- | --- |
|  | **Unvaccinated** | **Vaccinated** | **Unvaccinated** | **Vaccinated** |
| **Children (0-19)** | 679 147 | 428 207 | 4051 | 1555 |
| **Adults (20+)** | 1 213 027 | 2 476 374 | 9757 | 20 143 |
| **All cases** | 1 892 173 | 2 904 581 | 13 808 | 21 698 |
| **Total** | 4 832 260 | | | |

^a^ Of hospitalised cases, 92% are ward only, while 8% include an ICU stay.^24^ Ward vs. ICU classification is needed for acute COVID-19 morbidity estimates but is not applied for long COVID morbidity.

Data sources: covid19live.com.au, ABC case report, NSW archived surveillance reports.^26,32,34,35^

### **Supplementary Table 6: Omicron wave deaths by age**

| **Age group** | **Number of deaths** | **Percentage of deaths by age group** |
| --- | --- | --- |
| **0-10** | 7 | 0.19% |
| **11-19** | 3 | 0.09% |
| **20-29** | 8 | 0.24% |
| **30-39** | 34 | 0.98% |
| **40-49** | 56 | 1.62% |
| **50-59** | 122 | 3.52% |
| **60-69** | 332 | 9.59% |
| **70-79** | 736 | 21.25% |
| **80+** | 2165 | 62.52% |
| **All cases** | 3463 | |

Note that deaths are from reporting period December 15^th^-April 10^th^.

Data source: COVID-19 National Surveillance Epidemiology Report.^30^

### **Supplementary Table 7: Long COVID morbidity expected for any COVID-19 case (where morbidity is the proportionate loss in quality of life over 1 year compared to full health)**

| **Population group** | | **Morbidity estimate (95% UI)** | |
| --- | --- | --- | --- |
|  |  | **Unvaccinated** | **Vaccinated** |
| **Adults** | **Non-hospitalised** | 0.0016 (0.0007-0.0026) | 0.0009 (0.0004-0.0014) |
|  | **Hospitalised** | 0.0466 (0.0192-0.0741) | 0.0198 (0.0082-0.0314) |
| **Children** | | 0.0003 (0.0001-0.0004) | 0.0001 (0.0001-0.0002) |

Note the morbidity loss for someone actually with long COVID is much greater than shown here, as these are estimates of expected morbidity loss for any surviving COVID-19 case (where ‘case’ defines those who are symptomatic during the acute infection).

95% UI estimated using +/- 30% standard deviation.

UI= Uncertainty Interval

### **Supplementary Table 8: long COVID morbidity parameter study comparison**

| **Study** | **Symptoms considered** | **Health state** | **Morbidity ^a^** |
| --- | --- | --- | --- |
| Wulf Hanson et al. ^20^ (three levels of long COVID severity, across all COVID-19 cases) | Fatigue  Cognitive problems (mild)  Shortness of breath (mild) | Post-acute consequences (characterised by fatigue, muscle/joint pain, depression) (DW=0.219)  Dementia, mild (DW=0.069)  Chronic respiratory problems, mild (DW=0.019) | 0.00035 |
|  | Fatigue  Cognitive problems (severe)  Shortness of breath (moderate) | Post-acute consequences (characterised by fatigue, muscle/joint pain, depression) (DW=0.219)  Dementia, moderate (DW=0.377)  Chronic respiratory problems, moderate (DW=0.019) | 0.00084 |
|  | Fatigue  Cognitive problems (severe)  Shortness of breath (severe) | Post-acute consequences (characterised by fatigue, muscle/joint pain, depression) (DW=0.219)  Dementia, moderate (DW=0.377)  Chronic respiratory problems, moderate (DW=0.019) | 0.00107 |
| Our estimate (one level of long COVID severity, for community COVID-19 cases) | Total symptom profile considered per main analysis | Health states applied per main analysis | 0.0009 |
|  | Only including symptoms equivalent to health states applied by Wulf Hanson et al. (above): fatigue ^b^, cognitive problems, shortness of breath | Fatigue (DW=0.051)  Muscle/joint pain (DW=0.023)  Concentration difficulty (DW=0.069)  Chronic respiratory problems, mild (DW=0.019) | 0.00043 |

^a^ Morbidity per previously symptomatic acute COVID-19 survivor, each multiplied by OR=0.25 and OR=0.55 to reflect the morbidity among vaccinated individuals infected with the Omicron variant, as per our main methods.

^b^ The IHME applied ‘post-acute consequences’ to represent fatigue; this health state incorporates fatigue, as well as body pain, and depression. Therefore, in the comparison to our study, the health states we applied to represent both fatigue, and muscle/joint pain, were applied to more accurately compare to ‘post-acute consequences’ (noting that depression was not included in our main analysis for community cases so was not included here).

DW= Disability Weight.

### **Supplementary Table 9: long COVID burden of disease study comparison**

| **Study** | **DW (health state)** | **Duration** | **Prevalence ^a^** | **Morbidity estimate** | **Adjusted morbidity estimate ^b^** |
| --- | --- | --- | --- | --- | --- |
| **Wyper et al. ^38 c^** | 0.219 (post-acute consequences | 28 days | 14.3% | 0.0024 | 0.00033 |
| **Cuschieri et al. ^39 c,d^** | 0.219 (post-acute consequences | 28 days | 14.3% | 0.0024 | 0.00033 |
| **Moran et al. ^40^** | 0.219 (post-acute consequences | 28 days | 13.3% | 0.0022 | 0.00031 |
| **AIHW ^10^** | 0.219 (post-acute consequences | 91 days | 13.7% (adjusted down to represent only those with at least moderate impairment from long COVID) | 0.0045 | 0.00062 |
| **Wulf Hanson et al. ^20^** | 0.219 (post-acute consequences | 4 months | 18.1% (Fatigue)  10.1% (cognitive symptoms)  33.3% (respiratory symptoms ^e^ | 0.0025 | 0.00035 |
| **Our estimate** | See main results | | | | 0.0009 |

### ^a^ In each study, prevalence is measured among COVID-19 survivors.

^b^ Adjusted for vaccination and Omicron variant, per main analysis.

^c^ Same research group for ^38,39^.

^d^ Multiple sensitivity analyses run varying each parameter (duration increased, occurrence rate increased, parameters only applied to symptomatic cases).

^e^ Co-occurrence of each symptom pair, and all three symptoms together, was also measured.^20^

### **Supplementary Table 10: varying the risk of long COVID among Omicron-infected, hospitalised patients**

| **Sub group** | | **Primary analysis YLDs (95% UI)** | **Sensitivity analysis ^a^ YLDs (95% UI)** | **Percentage difference** |
| --- | --- | --- | --- | --- |
| Unvaccinated | Hospitalised adults | 455 (178-684)) | 74 (30-117) | -84% |
|  | Community adults | 1967 (810-3123) | N/A | N/A |
|  | Children | 171 (70-271) | N/A | N/A |
| Vaccinated | Hospitalised adults | 399 (154-595) | 82 (34-131) | -79% |
|  | Community adults | 2197 (905-3488) | N/A | N/A |
|  | Children | 59 (24-94) | N/A | N/A |
| **Total YLDs ^b^** | | **5200 (2200-8300)** | **4600 (1900-7200)** | **-12%** |

^a^ OR of 0.25 applied to morbidity estimate for hospitalised base case group (as well as for community adult and child strata, as per main analysis), to achieve morbidity among Omicron-infected cases.

^b^ Note: raw data shown for sub-groups; rounded estimates shown for total YLDs.

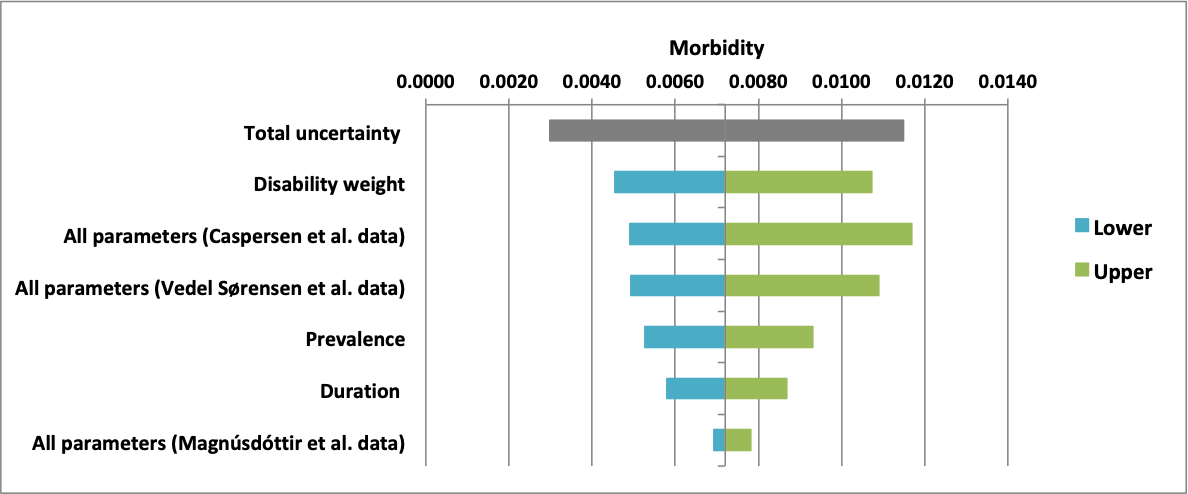

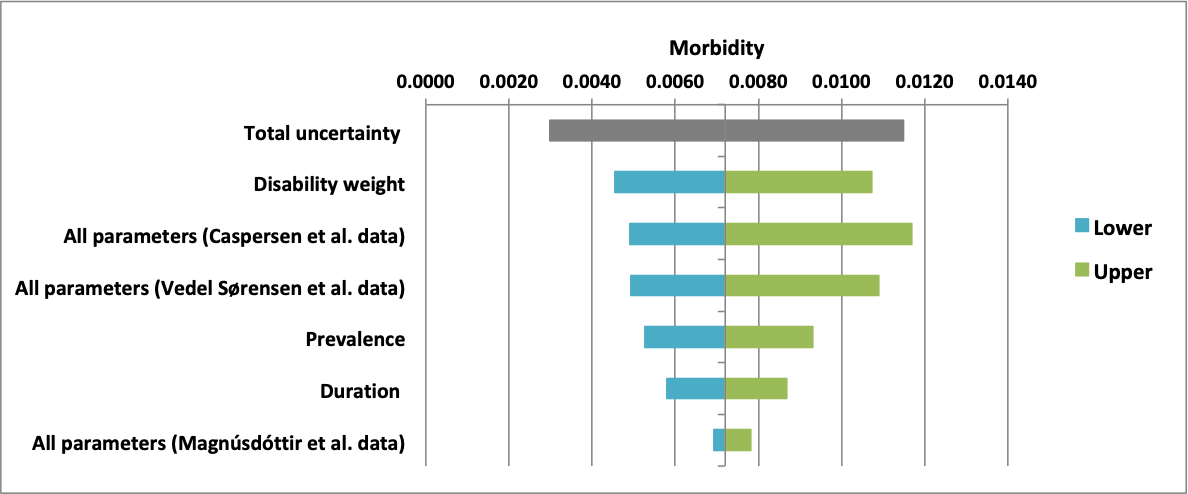

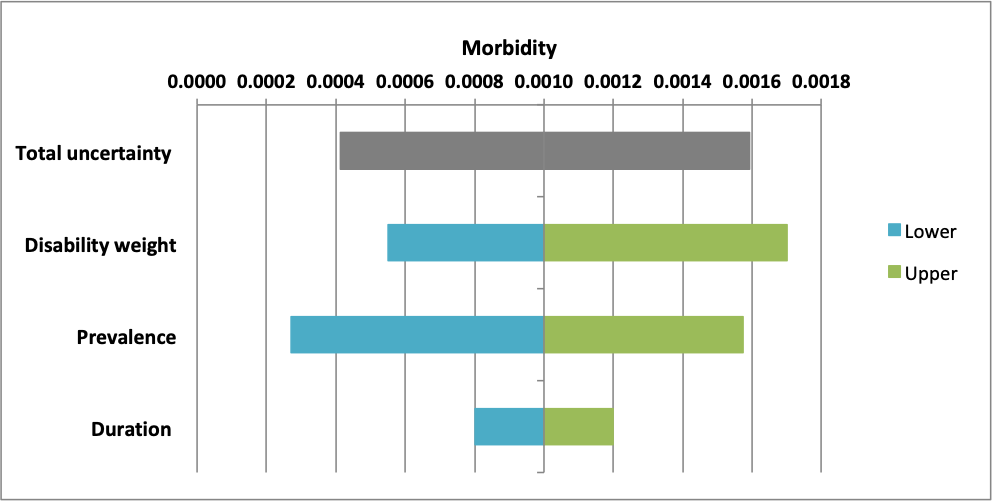

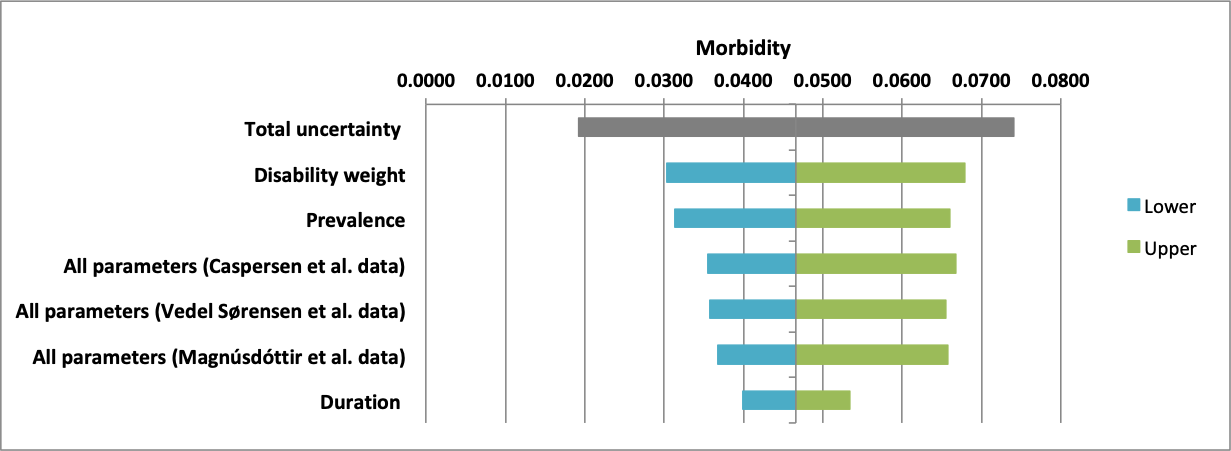

**A) Non-hospitalised adults (**$\boldsymbol{\geq}$**18)**

**B) Hospitalised adults (**$\boldsymbol{\geq}$**18)**

**C) Children (**$\boldsymbol{<}$**18)**

**Supplementary Figure 1: Tornado plots of base case morbidity estimate parameters and their contribution towards uncertainty in estimates.**

**Panel A:** Adults not hospitalised during acute infection. **Panel B:** Adults hospitalised during acute infection. **Panel C:** Children. All groups represent pre-Omicron variant, unvaccinated cases. The grey bars indicate the 95% uncertainty interval applied to each overall morbidity estimate (using +/- 20% standard deviation). Each parameter is varied individually (disability weight, prevalence, and duration) and also varied simultaneously by the source study of prevalence estimates (Sørensen et al., Caspersen et al., Magnúsdóttir et al.) for the adult sub-groups (a & b).^3,12,13^
